# Rethinking Depression in Cities: Evidence and Theory for Lower Rates in Larger Urban Areas

**DOI:** 10.1101/2020.08.20.20179036

**Authors:** Andrew J. Stier, Kathryn E. Schertz, Nak Won Rim, Carlos Cardenas-Iniguez, Benjamin B. Lahey, Luís M. A. Bettencourt, Marc G. Berman

**Author notes:** To whom correspondence should be addressed; and/or.

## Abstract

It is commonly assumed that cities are detrimental to mental health. However, the evidence remains inconsistent and, at most, makes the case for differences between rural and urban environments as a whole. Here, we propose a model of depression driven by an individual’s accumulated experience mediated by social networks. The connection between observed systematic variations in socioeconomic networks and built environments with city size provides a link between urbanization and mental health. Surprisingly, this model predicts lower depression rates in larger cities. We confirm this prediction for US cities using three independent datasets. These results are consistent with other behaviors associated with denser socioeconomic networks and suggest that larger cities provide a buffer against depression. This approach introduces a systematic framework for conceptualizing and modeling mental health in complex physical and social networks, producing testable predictions for environmental and social determinants of mental health also applicable to other psychopathologies.

---

Living in cities changes the way we behave and think (*1, 2*). Over a century ago, the social changes associated with massive urbanization in Europe and in the United States, focused social scientists on the nexus between cities and mental life (*2*). Along with urban public health crises of the time, a central question became whether cities are good or bad for mental health.

Subsequently, social psychologists (*1*) started to document and measure the systematic behavioral adaptations among people living in cities. These adaptations included strategies to curb unwanted social interactions – such that people in larger cities act in colder and more callous ways (*1*), a more intense use of time (e.g. faster walking (*3*)), and a greater tolerance for diversity (*4*). These studies attributed the influences of urban environments on mental health to the intensity of social life in larger cities, mediated by densely built spaces and associated dynamic and diverse socioeconomic interaction networks. They did not, however, ultimately clarify whether urban environments promote better or worse mental health. Consequently, concerns persisted that cities are mentally taxing (*5–8*) and can induce “stimulus overload”, including stress and mental fatigue (*9*).

More recent studies have focused less on urban environments as a whole and more on contextual and environmental factors affecting individuals. For example, a study of the entire population in Sweden (*8*) uncovered a positive association between neighborhood population density and depression-related hospitalizations. In addition, individual factors of gender, age, socioeconomic status, and race, which vary at neighborhood levels within cities, have been found to be statistically associated with depression (*10–12*). Other studies using various measures of mental health and broader definitions of urban environments have found evidence for an association between poorer mental health in cities versus rural areas (*6, 7*). However, this evidence has remained mixed and often explicitly inconsistent (*13,14*), due to differences in: 1) reporting (e.g., surveys vs. medical records), 2) types of measurement (e.g., surveys vs. interviews), 3) definitions of what constitutes urban, and 4) the mental disorders studied (e.g. schizophrenia vs. depression).

For these reasons, it is desirable to create a systematic framework that organizes this diverse body of research and interrogates how varying levels of urbanization influence mental health across different sets of indicators. Here, we begin to build this framework for depression in US cities. We show that, surprisingly, the per capita prevalence of depression decreases systematically with city size.

Like earlier classical approaches, our strategy frames the effects of city size on mental health through the lens of the individual experience of urban physical and socioeconomic environments. Crucial to our purposes, many characteristics of cities have been recently found to vary predictably with city population size. These systematic variations in urban indicators are explained by denser built environments and their associated increases in the intensity of human interactions and resulting adaptive behaviors (*15*).

More specifically, people in larger cities have, on average, more socioeconomic connections, mediating a greater variety of functions. This effect is understood in theoretical terms by the statistical likelihood to interact with more people in space, leading to both potential mental “overload” but also to greater stimulation and choice along more dimensions of life. This expansion of socioeconomic networks is supported structurally by economies of scale (e.g., road length) in urban built environments and by individual occupation specialization and associated increases in economic productivity and exchange.

This effect leads to a number of quantitative predictions about the nature of urban spaces and socioeconomic variables, the most central of which is the variation of the average number of socioeconomic interactions, *k* (network degree) with city size, *N*, as *k*(*N*) = *k*_0_*N^δ^*. Here, *k*_0_ is a prefactor independent of city size, and *ξ* a residual measuring the distance from the population average. The exponent 0 < *δ* ≃ 1/6 < 1 measures the percent increase in the number of connections with each percent increase in city population, which is an elasticity in the language of economics. Because the *ξ* reflects city-size independent statistical fluctuations, these errors average out across cities and *k* obeys a scaling relationship on average over cities, such that *k*(*N*) ∼ *N^δ^*. This expectation is directly observed in cell phone networks (*16*) and indirectly via the faster spread of infectious diseases such as COVID-19 (*17*), and by higher per capita economic productivity and rates of innovation (*3, 15*).

This result is important to mental health because depression is associated, at the individual level, with fewer social contacts (*18, 19*). To translate the general scaling of social interactions with city size into a model for the incidence of depression in urban areas, we will now need to pay particular attention not only to the average number of social connections in a city of size *N, k*(*N)*, but also to its variance across individuals in that city and how they influence depression.

To do this, we developed a statistical mathematical model that combines socioeconomic network structure with individual risk of depression (Figure 1). This model takes the form of a generative social network, which combines: i) a degree distribution with mean scaling as *k*(*N*) = *k*_0_*N^δ^* (Figure 1B) with ii) the risk (probability) for an individual to manifest depression, *p_d_*(*k*), taken to be inversely proportional to their social connectivity, *p_d_*(*k*) ∼ 1/*k* (Figure 1C). We will return to the finer issue of quality and type of connections below. For now, note that a larger number of connections in larger cities entails a qualitatively different experience because it is driven by the need to obtain support, goods, and services in environments with deep divisions of knowledge and labor.

**Figure 1:**
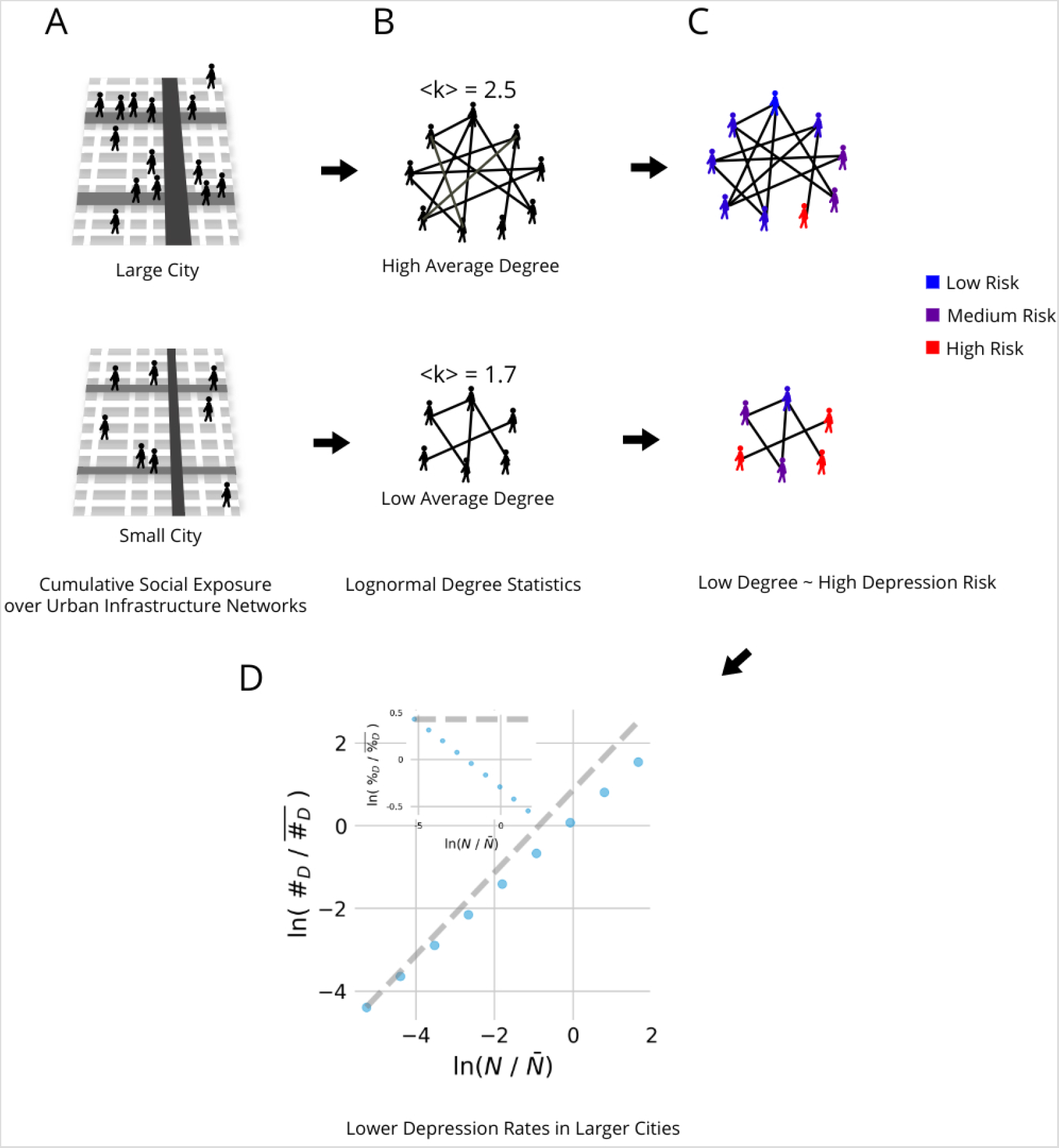
Sub-linear scaling of depression in a social network model. (a) Individuals moving over a city’s hierarchical infrastructure network experience cumulative exposure to semirandom social interactions. (b) This cumulative exposure results in social networks with logskew-normal degree (k) statistics with a mean which increases with city size indicating more per-capita social interactions in larger cities, on average. (c) Individual risk for depression is inversely proportional to social connectivity (degree) and is superimposed on the social networks generated within cities (d) The combination of how cities shape social networks and how social networks shape individual depression risk results in a prediction of sub-linear scaling of depression cases with increased city size, i.e., lower depression rates in larger cities (Inset). The logarithm of population and depression incidence are mean centered for ease of comparison to the empirical results.

To complete the model, we need to specify the probability distribution of degree, *f(k)* in each city. We adopt a log-skew-normal distribution with parameters similar to those measured in (*16*), see Figure 1B. This choice introduces another assumption into our model because lognormal distributions arise from multiplicative random processes, which compound risk over time to generate outcomes. In this sense, the adoption of this distribution assumes that depression is the result of a cumulative exposure process over time (*20*), see Figure 1A, mediated by an individual’s social network. Figure 1D shows results from this model obtained by sampling each city’s degree distribution *N* times, corresponding to a city’s population. Each simulated city resident is then diagnosed with a binary outcome, manifesting depression or not proportionally to their individual risk, *p_d_*(*k*).

We used this model to generate urban socioeconomic networks and computed their associated number of depression cases, *Y*, for a range of city sizes from *N* = 10^4^ −10^7^ that span population sizes for US metropolitan areas, see Figure 1D. We observed a simple scaling relation for the total number of depressive cases

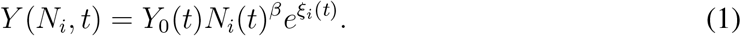

with a sub-linear exponent *β* = 1 − *δ <* 1. For *β* = 1 (*δ* = 0), cases of depression increase proportionally to population so that there would be no city size effect. In contrast, for *β <*1 (sub-linear), a smaller proportion of the population manifests depression in larger cities.

We express the quantitative consequences of the model based on 100 iterations for each city to predict that the number of depression cases follows a power law function of city size with a scaling exponent *β* = 0.859 (95% CI = [0.854, 0.863]), Figure 1D. Thus, under the model’s assumptions, we expect larger cities to show substantially lower per capita rates of depression.

To test these quantitative expectations, we asked whether empirical measurements of depression exhibit a systematic scaling relationship with city population size. We analyzed three new data sets, which allow for consistent assessments of cases of depression across different urban areas in the US.

First, we employed estimates of the prevalence of depression in US cities produced as a part of two annual population surveys: the National Survey on Drug Use and Health (NSDUH) (*21*) from the Substance Abuse and Mental Health Services Administration (SAMHSA) and the Behavioral Risk Factor Surveillance System (BRFSS) (*22*) from the Centers of Disease Control (CDC), see Supplementary Text, Supplementary Figures 1, 2, and Supplementary Tables 4,2,3. The NSDUH asks respondents whether they have experienced a major depressive episode in the past year, as defined by the *Diagnostic and Statistical Manual of Mental Disorders* (DSMIV) (*21*). The BRFSS asks respondents if they have ever been told that they have a depressive disorder. Both surveys involved a social interaction between a surveyor and the respondent, which takes place over the phone for the BRFSS and in person for the NSDUH. The differences between the two surveys provide a consistency test on measured cases of depression and partially rule out the possibility that their variation with city size is idiosyncratic to particular experimental or survey methodologies.

Second, to generalize across different indicators and to avoid biases in reporting due to social stigma (*23*), we added an additional estimate of depression prevalence based on passive observation, which does not rely on an overt survey instrument. Specifically, we explored a large geo-located Twitter dataset of individuals and their messages for depressive symptoms in different cities over one week in 2010 (*24*). Similar datasets have been used to demonstrate that happiness decreases with per-capita tweets (*25*) and that counts of users scale super-linearly with city size (*26*), but to our knowledge, they have not been used to directly estimate associations between mental health disorders and city size.

To measure the prevalence of depression from this corpus, we employed a machine learning technique to identify depressive symptoms from users’ messages, emulating the Patient Health Questionnaire-9 (PHQ-9) commonly used by clinicians. The PHQ-9 consists of 9 questions based on the nine criteria for diagnosing depression in the DSM-IV. In order to emulate the PHQ-9 questions, we used a previously determined lexicon of seed terms organized into nine topics to guide a Latent Dirichlet Allocation (*27*) method to determine the degree to which each user’s messages represent these topics, see Supplementary Text, Supplementary Figure 3, and Supplementary Table 5. This technique has been found to have an accuracy (proportion of tweets correctly identified) of 68% and precision (1 the false discovery rate of tweets with depressive symptoms) of 72% compared to expert assignment of tweets to PHQ-9 questions (*27*).

**Figure 2:**
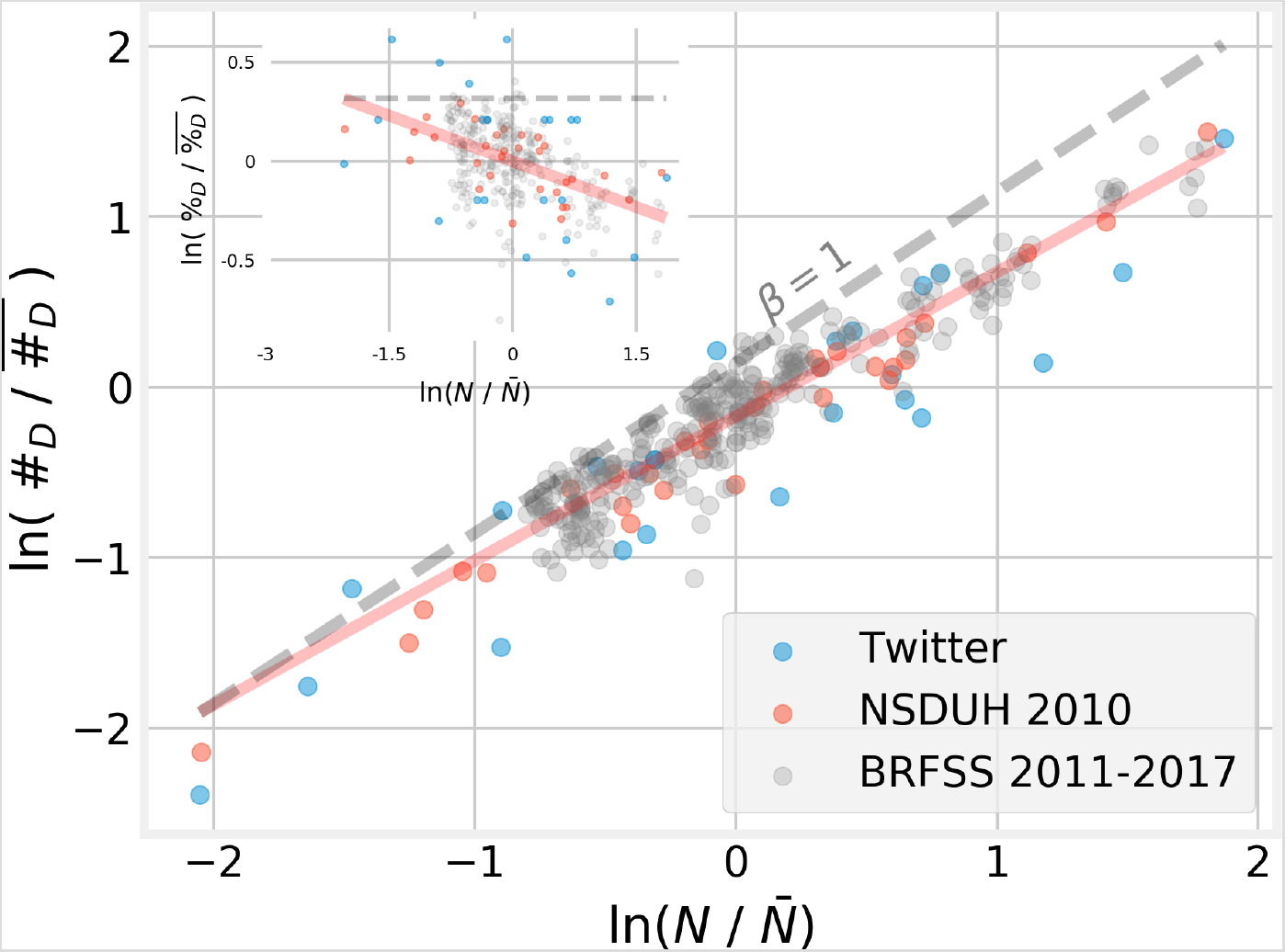
Depression cases scale sub-linearly with city size. City level measures of depression prevalence were obtained from two survey based data sets (NSDUH and BRFSS) and one passive observation data set (Twitter). To collapse across datasets the natural log of Population, *N*, and estimated total depression cases, #_*D*_, were mean-centered within each dataset. An ordinary least squares linear regression of the pooled data resulted in an estimate of *β* = 0.847, 95% CI = [0.816, 0.879], and an *R*^2^ of 0.89. Inset: depression rates decrease with city size. *β* = −0.15, 95% CI = [-0.18, –0.12], *R*^2^ = 0.23.

We estimated the scaling exponent *β* from each of these datasets via ordinary least squares (OLS) linear regression between the logarithm of total depression cases and the logarithm of population size (See Supplementary Text, Supplementary Figures 4, 5, Supplementary Table 6). When pooling across datasets and years we estimated a scaling exponent of *β* = 0.847 (95% Confidence Interval (CI) = [0.815, 0.878]) (Figure 2), consistent with our simulation model’s prediction of *β* = 0.859. Moreover, estimates of *β* are similar when calculated separately for each dataset (Table 1). This statistical relationship between depression and city size is consistent across all three datasets despite the different ways in which depressive symptoms are measured and the different ways that the data were collected. Importantly, these results demonstrate that depression rates are substantially lower in larger US cities, contrary to previous expectations, but precisely inline with our theoretical model and simulations.

**Table 1:**
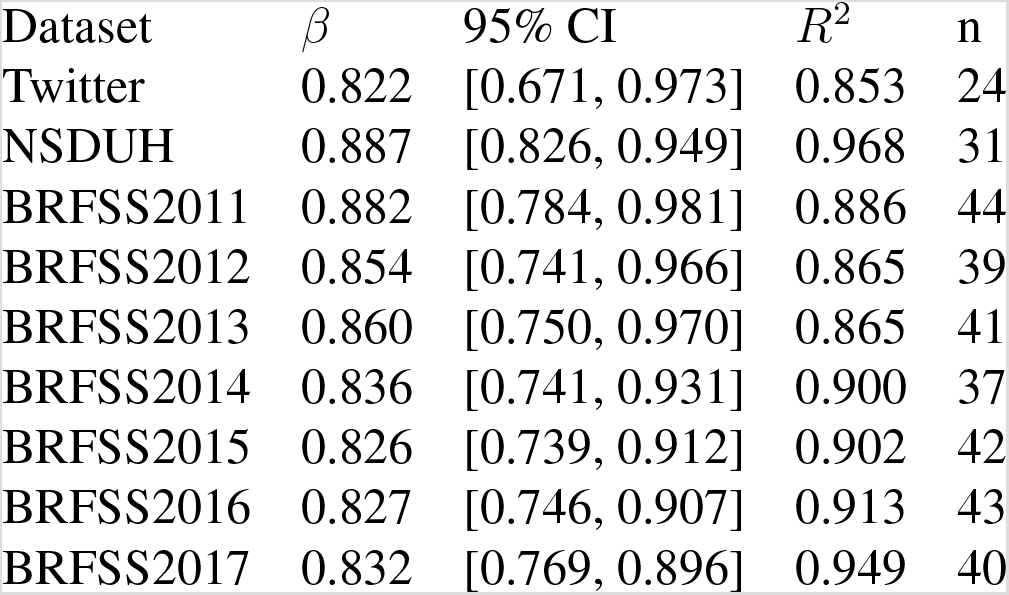
Estimates of the scaling exponent *β* for each dataset. In all cases we observe sub-linear (*β <*1) scaling of total depression cases with city size. *n* indicates the number of cities included in each dataset.

| Dataset | $\beta$ | 95% CI | $R^2$ | n |
| --- | --- | --- | --- | --- |
| Twitter | 0.822 | [0.671, 0.973] | 0.853 | 24 |
| NSDUH | 0.887 | [0.826, 0.949] | 0.968 | 31 |
| BRFSS2011 | 0.882 | [0.784, 0.981] | 0.886 | 44 |
| BRFSS2012 | 0.854 | [0.741, 0.966] | 0.865 | 39 |
| BRFSS2013 | 0.860 | [0.750, 0.970] | 0.865 | 41 |
| BRFSS2014 | 0.836 | [0.741, 0.931] | 0.900 | 37 |
| BRFSS2015 | 0.826 | [0.739, 0.912] | 0.902 | 42 |
| BRFSS2016 | 0.827 | [0.746, 0.907] | 0.913 | 43 |
| BRFSS2017 | 0.832 | [0.769, 0.896] | 0.949 | 40 |

In summary, although the association between urbanization and mental health is foundational in the social sciences and in public health, it has remained challenging to characterize and assess quantitatively. This is particularly concerning as almost every nation worldwide continues to urbanize, with over 70% of the world’s population expected to live in cities by 2050 (*28*), and depressive disorders already a leading global cause of disability (*29*) and economic losses (*30*).

The convergence of recent findings from urban science with evidence and theory from mental health studies offers a new window for creating more systematic approaches to understanding mental health in cities. In this respect, the sub-linear scaling of total depression cases with population size is a completely unexpected result characterizing the socio-geographic distribution of depression. It suggests that larger city environments and urbanization may, on average, actually provide greater social stimulation and connections that may buffer against depression.

At the same time, we must recognize that the numerous factors which may influence depression vary enormously within cities. These variations may influence individuals directly and also indirectly through the local environments in which they live and work. Looking within cities at these local and more fine-grained levels is expected to reveal variations in the incidence of depression by neighborhood and via other social groupings (*10*). For example, several studies have associated high population density in social housing in Europe and the US with higher incidence of depression in aging adults (*13*), possibly mediated by a higher density of negative connections with neighbors, which can instill feelings of isolation, fear and despair.

More generally, neighborhood level tracking of depression under conditions of strong sorting or segregation is likely to reveal signatures of social network determinants of depression that may interact with local variations of other individual risk factors, such as age, race and socioeconomic status. Future work may employ a number of experimental designs such as sibling comparisons or stratification by confounding factors to search for finer causal evidence (*31*) for the influence of social networks on depression.

Examining scaling relationships of mental health outcomes with city size is a novel and systematic way of investigating general urban effects on mental life. The perspective of cities as interconnected networks which shape their inhabitants lives may also help to uncover environmental factors that influence other mental health disorders. This includes highly co-morbid psychopathologies such as anxiety disorders, and less co-morbid ones such as schizophrenia, for which increased socialization may lead to different outcomes. The fact that important insights about the mechanisms of mental health disorders might be gleaned from such a general population level analysis, which ignores the intricate and often personal details of mental health, is surprising and powerful.

## Data Availability

All data are publicly available and all relevant code is linked in the manuscript

## Acknowledgments

Conceptualization, A.J.S., L.M.A.B. and M.G.B.; Methodology, A.J.S. and M.G.B.; Formal Analysis, A.J.S; Investigation, A.J.S, M.G.B., and N.W.R.; Data Curation, A.J.S., K.E.S., C.C.I., N.W.R., and M.G.B.; Writing – Original Draft, A.J.S., M.G.B., and L.M.A.B; Writing – Review & Editing, A.J.S., M.G.B., L.M.A.B., B.B.L., K.E.S., N.W.R., and C.C.I.; Supervision, M.G.B. and L.M.A.B. This work was partially supported by NSF grants DGE-1746045 to K.E.S. BCS-1632445 and SCC-1952050 to M.G.B.

## Supplementary Materials

### Materials and Methods

#### Data Sources and Processing

County populations in Figure 2 are provided by the United States Census Bureau and available online at https://www.census.gov/data/tables/time-series/demo/popest/2010s-counties-total.html#par_textimage. We used delineation files provided by the US Office of Budget and Management to roll up county level data to Metropolitan Statistical Areas (MSAs). Each MSA is represents a US Census definition of a functional city in the USA, circumscribing together a city and its suburbs, sometimes known as an integrated labor market in economic geography. These definitions are updated regularly and available https://www.census.gov/programs-surveys/metro-micro/about/delineation-files.html. The list of MSA included in analysis in the main text (Figure 2) are enumerated in Supplementary Table 1.

As the surveys from which we obtained depression prevalence estimates are administered by different agencies (the Substance Abuse and Mental Health Services Administration administers the NSDUH and the Centers for Disease Control and Prevention administers the BRFSS), collection and reporting methods differ substantially between these two data sources. Importantly, the NSDUH is conducted in person while the BRFSS is conducted over the phone. In addition, the two surveys differ in the questions they ask about depressive symptoms. The NSDUH asks participants whether or not they had a period of 2 or more weeks in which they experienced depressive symptoms in line with definitions in the DSM-IV (*32*). In contrast, the BRFSS asks respondents if they have ever been told that they “have a depressive disorder (including depression, major depression, dysthymia, or minor depression)?” (*22*). In addition to these differences in questionnaire content and methods, the two data sources also differ in how they report data. The NSDUH reports age, ethnicity, and geography adjusted prevalence estimates in 33 metropolitan statistical areas (MSAs) (*21*). In contrast, the BRFSS reports age, gender, and socioeconomically adjusted prevalence estimates for any MSA with at least 500 respondents (*22*), and consequently the cities which are included in reports vary from year to year.

The 2010 NSDUH estimates of the rate of major depressive episodes used in Figure 2 were obtained from Table 38 of the Substance and Mental Health Services Administration 2005 - 2010 National Survey on Drug Use and Health. This data are available online at https://www.samhsa.gov/data/sites/default/files/NSDUHMetroBriefReports/NSDUHMetroBriefReports/NSDUH_Metro_Tables.pdf. We multiplied estimated prevalence by 2010 estimated population to determine the estimate of total depression cases within each MSA.

The 2011–2017 BRFSS city estimates of the prevalence of major depression used in Figure 2 are available online at https://www.cdc.gov/brfss/smart/Smart_data.htm. As with the NSDUH data we multiplied estimated prevalence by that years estimated population to estimate of total depression cases within each MSA.

One point of concern was that the cutoff of 500 respondents per city in the BRFSS data might artificially alter the joint distribution of prevalence estimates and city size (i.e which cities are included in the sample) in a way that biases the estimate of *β*, the slope of the scaling line. One possibility is that larger cities are simply more likely to record enough responses to be included. However, since the BRFFS data includes cities with populations as small as 20,285, whatever bias this 500 respondent cuttoff may introduce likely has a more complex origin. In order to address this without knowing the source of potential biases in city inclusion we employed non-parametric change point detection based on the minimum covariate discriminant (MCD) (*33*) in order to find the city size at which the joint distribution of city size and depression prevalence was different on either side of the change point. This was applied to each each year of BRFSS data separately and results were consistent across years with the mean change point of (692,557 people, sd = 268,004 people).

Specifically, we followed a procedure similar to (*34*). For each year of BRFSS data we first ordered the data by population and then applied a python implementation of the MCD algorithm (*35*) with a sliding window. This resulted in a robust-to-outliers estimate of the mean within the window and the 2-by-2 robust covariance matrix between population and depression prevalence within the window. These two quantities allow for the estimation of the Mahalanobis distance between the robust mean and the data from the city which has the next smallest population to the smallest city included in the window (the left out city). These distances follow a chisquared distribution with degrees of freedom equal to the size of the window. Consequently we marked the left out city as a potential change point if the Mahalanobis distance was greater than the 97.5th percentile of the relevant chi-squared distribution. Finally we calculated a moving average with window size five, of marked change points. We considered a specific city size to be a change point if the moving average of marked change points was greater than 0.5. This was repeated for MCD window sizes from 5 to 25 data points in increments of 2. Histograms of the detected change points over all window sizes are shown in Supplementary Figure 1.

Next we used a k-means clustering implementation in python (*35*) to split the detected change points into two separate clusters based on the observation that the histograms of change points over all bin sizes for most years are roughly bimodally distributed. We used the two cluster centers as the final change points for each year of BRFSS data resulting in a partition of the data into three sets. Scaling estimates for the largest cities are reported in the main text and Figure 2 and Table 1. When pooling all BRFSS data across all cities and years we still find evidence that larger cities have lower depression rates than smaller cities *β* = 0.926 (95% CI = [0.903, 0.950]). Results are similar when *β*is estimated separately for each year of BRFSS data (Supplementary Table 4). When pooling data from the other two partitions which contain smaller cities, we found no evidence that depression rates scale sub- or super-linearly with population *β* = 0.996 (95% CI = [.956,1.035]) (Supplementary Figure 2). Results were similar when estimating *β*for each year separately (Supplementary Table 2). This lack of a city size effect for smaller cities in the BRFSS data may indicate that social network determinants of depression are overshadowed by other risk factor in smaller cities, but may also be specific to biases introduced by the way in which the data were collected and reported.

We further estimated the sensitivity of our *β* estimates among larger cities to variation in the change point. For each year of BRFSS data we varied the change point 100 times according to a normal distribution with a mean of the change point used for that year in the main text and a variance equal to the variance in change points across years. We found that the estimates of *β* in larger cities are robust to these variations in the choice of change point (Supplementary Table 3).

The geolocated Twitter dataset used in Figure 2 and Table 1 is available online at http://www.cs.cmu.edu/~ark/GeoText/. This dataset included 377,616 tweets from 9,475 users collected over a one week period in March of 2010 (*24*). Latitude and Longitude coordinates for each tweet were converted to a county-level Geographic Identifier (GEOID) using the US Census Geocoder API provided by the United States Census Bureau available at https://github.com/fitnr/censusgeocode. If there was more than one coordinate per user, we used the mode and in the case of a tie, we used the coordinate that appeared first in time. We then used delineation files provided by the US Office of Budget and Management to roll up county level data to MSAs.

We processed tweets following (*27*), using standard text preprocessing (for example, deleting stop words) and processing steps specific to the twitter platform (for example, deleting “#” in the hashtags). Then, we used a previously determined lexicon of seed terms related to depression symptoms organized into nine topics based on the PHQ-9, to guide a Latent Dirichlet Allocation (LDA) model (*27*). LDA allows for the discovery of underlying topics within collections of text data and has been utilized previously with short, semi-structured text sources (e.g. (*36, 37*)). This enabled us to find users who had topic cluster(s) related to nine PHQ-9 topics in their tweets over one week.

One point of concern was that individuals who have depressive symptoms may tweet differently from those that do not. Specifically, we worried that individuals with depressive symptoms would tweet less leading to less reliable estimates from these individuals. In order to control for this possibility we performed a logistic regression to predict the presence of depressive language in users tweets from their number of tweets over the 1 week collection period. We repeated this procedure excluding users who had fewer than a specified number of tweets for cutoffs from 0 tweets to 110 tweets. As demonstrated in Supplementary Figure 3, the logistic regression model achieves significance when individuals with fewer than 92 tweets are included. This indicates that people with depressive symptoms tend to tweet less, but that among individuals who tweeted at least 92 times over the collection period, a logistic regression model cannot differentiate between individuals with- and without- depressive symptoms based on their number of tweets. Consequently, we excluded individuals with fewer than 92 tweets and then estimated depression prevalence as the proportion of users in each city whose tweets contained a nonzero signal for any of the PHQ-9 topics. In addition, we excluded cities in which estimated depression rates were unrealist at 0% or 100%.

In order to test the sensitivity of the results to the minimum tweet threshold we repeated the scaling analysis on the Twitter dataset with minimum tweet count cutoffs from 82 to 101 (Supplementary Table 5) and found that estimates of *β* were robust to these changes in exclusion criteria.

## Code Availability

All relevant data processing code is available at https://github.com/enlberman/depression_scaling.

## Estimating the Scaling Exponent *β*

We performed OLS linear regression in order to calculate the scaling exponent *β*for depression cases. We verified that the residuals of the models in Table 1 are approximately normally distributed both with q-q plots of the residuals (Supplementary Figure 4) and the the Shapiro-Wilk test of normality (*38*) (Supplementary Table 6). We also verified that the residuals are not correlated with city size (Supplementary Figure 5, Spearman-r minimum p-value = .44).

## Simulating Depression In Cities

The starting point for the simulations of depression cases was the log-skew-normal degree distribution, which has been shown to match the degree distributions of cell phone based social networks in cities (*16*) and theoretically is the result of cumulative exposures to semi-random interactions taking place throughout cities’ infrastructure networks. The log-skew-normaldistribution has the density function

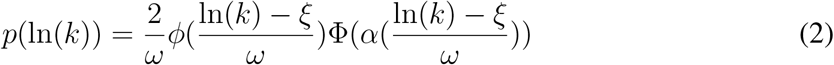

where *ξ* is the location parameter, *α* is the shape parameter, and *ω* is the scale parameter, *φ* is the normal distribution probability density function, and Φ is the normal distribution cumulative density function. These parameters can be transformed into the more familiar mean ( *µ*), variance ( *σ*^2^), and skewness (*γ*_1_) via (*39*):

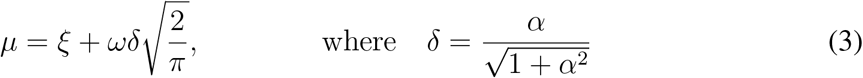

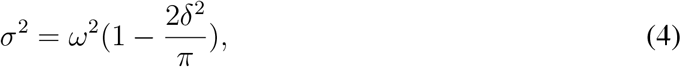

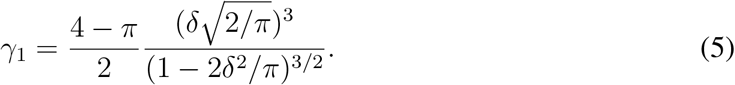

We started with values of *σ* = .87, *γ*_1_ = .2, and *µ* = 1 .97 in line with a city of size *N* = 10,000 (*16*). We then let the mean of this distribution grow with population size according to

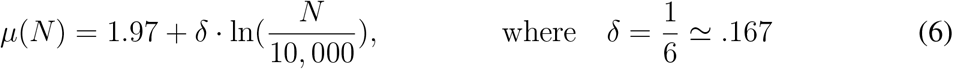

so that 〈*k*〉 ∼ *N^δ^*. For each simulated city with size, *N*, we sampled uniformly from it on a log scale from 10^4^ to 10^7^. We then sampled from the degree distribution *N* times to obtain a list of the social network degrees of all *N* simulated city inhabitants. From this list we randomly assigned each simulated individual to be diagnosed with depression (or not) with a probability inversely proportional to their degree (probability of depression ∼1/k). Total depression cases in each simulated city were calculated as the sum of depressed individuals.

## Supplementary Figures

**Supplementary Figure 1:**
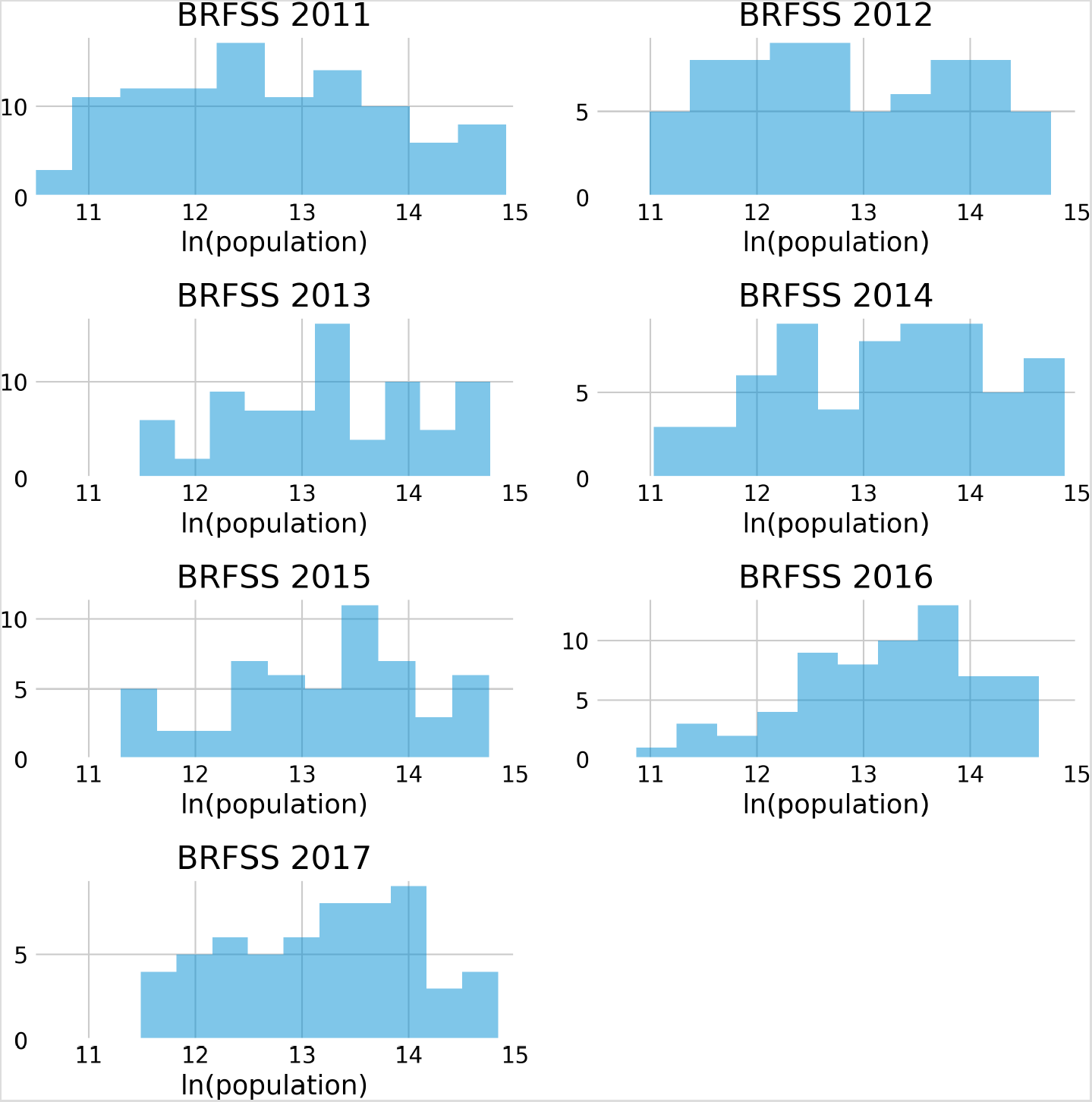
Histograms of the detected change points for all window sizes in BRFSS data.

**Supplementary Figure 2:**
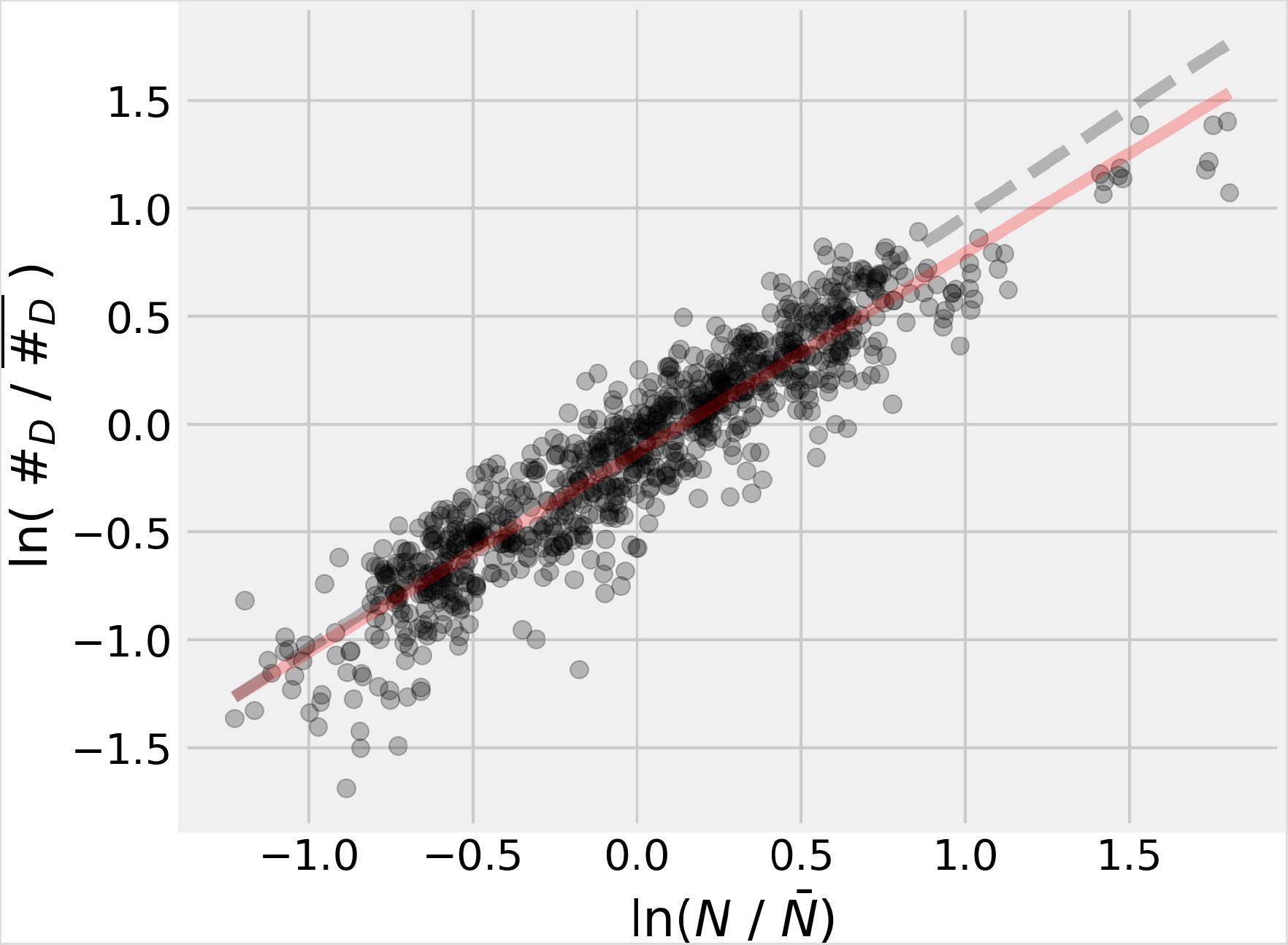
Pooling BRFSS data across years for all cities results in a scaling exponent of *β* = 0.926(95% CI = [0.903, 0.950]), consistent with lower depression rates in larger cities.

**Supplementary Figure 3:**
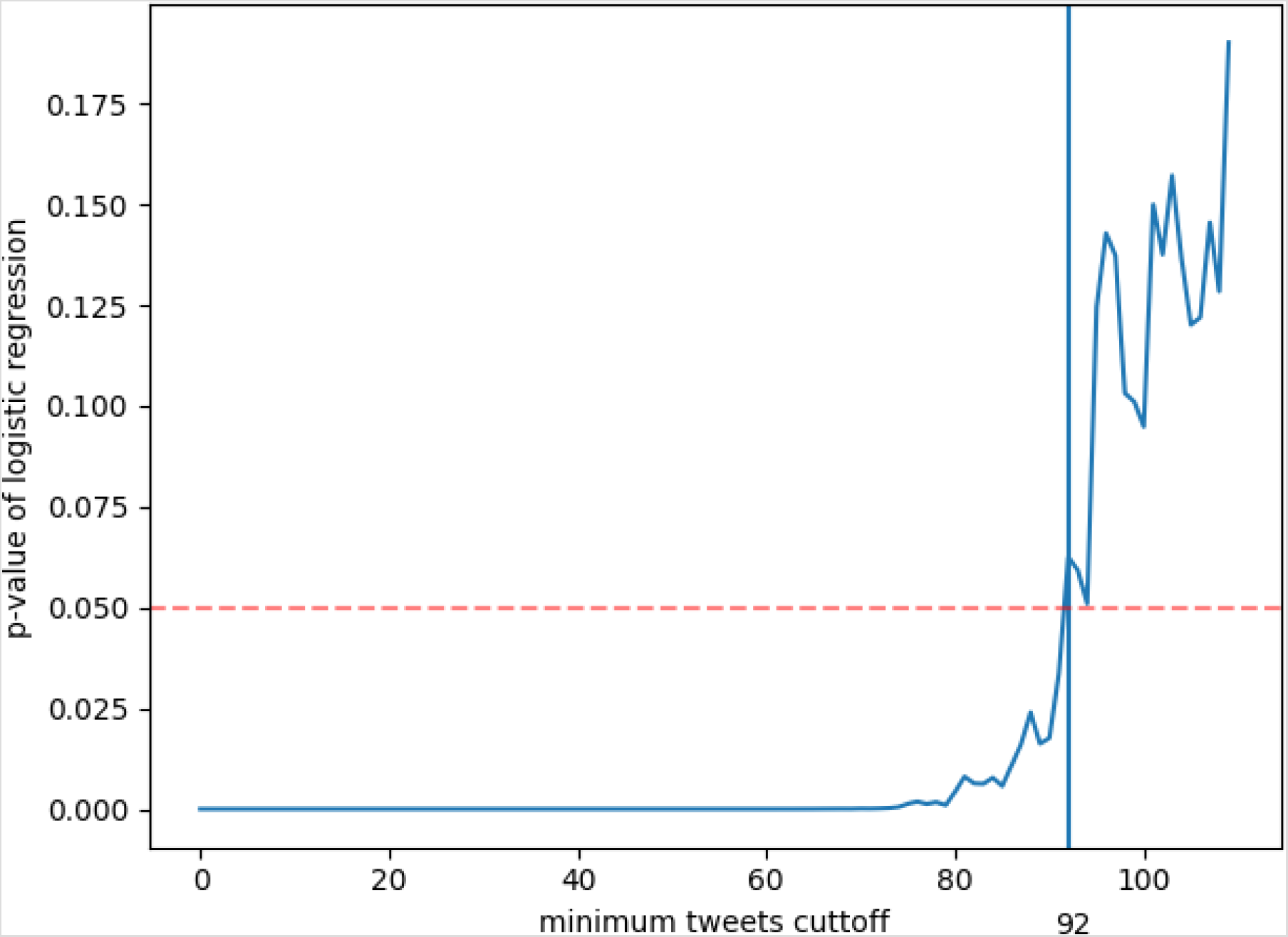
Users with lower numbers of tweets are more likely to have depressive sentiment in their tweets. When using an exclusion criteria of less that 92 tweets a logistic regression model significantly distinguishes individuals with depressive sentiment from individuals without depressive sentiment.

**Supplementary Figure 4:**
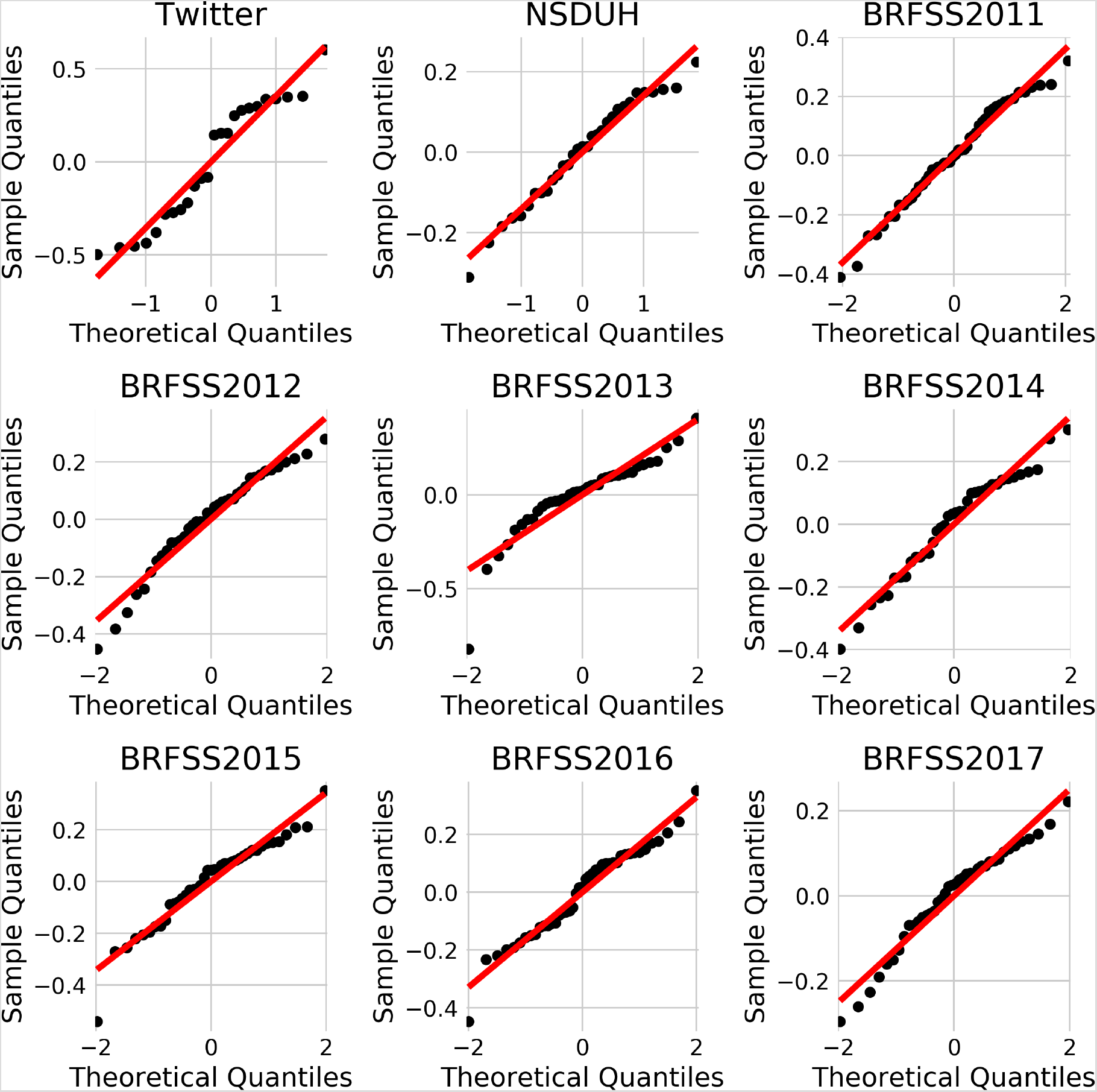
QQ plots of the residuals of the OLS model. No significant deviations are observed indicating that the residuals are approximately normally distributed and the linear model is appropriate.

**Supplementary Figure 5:**
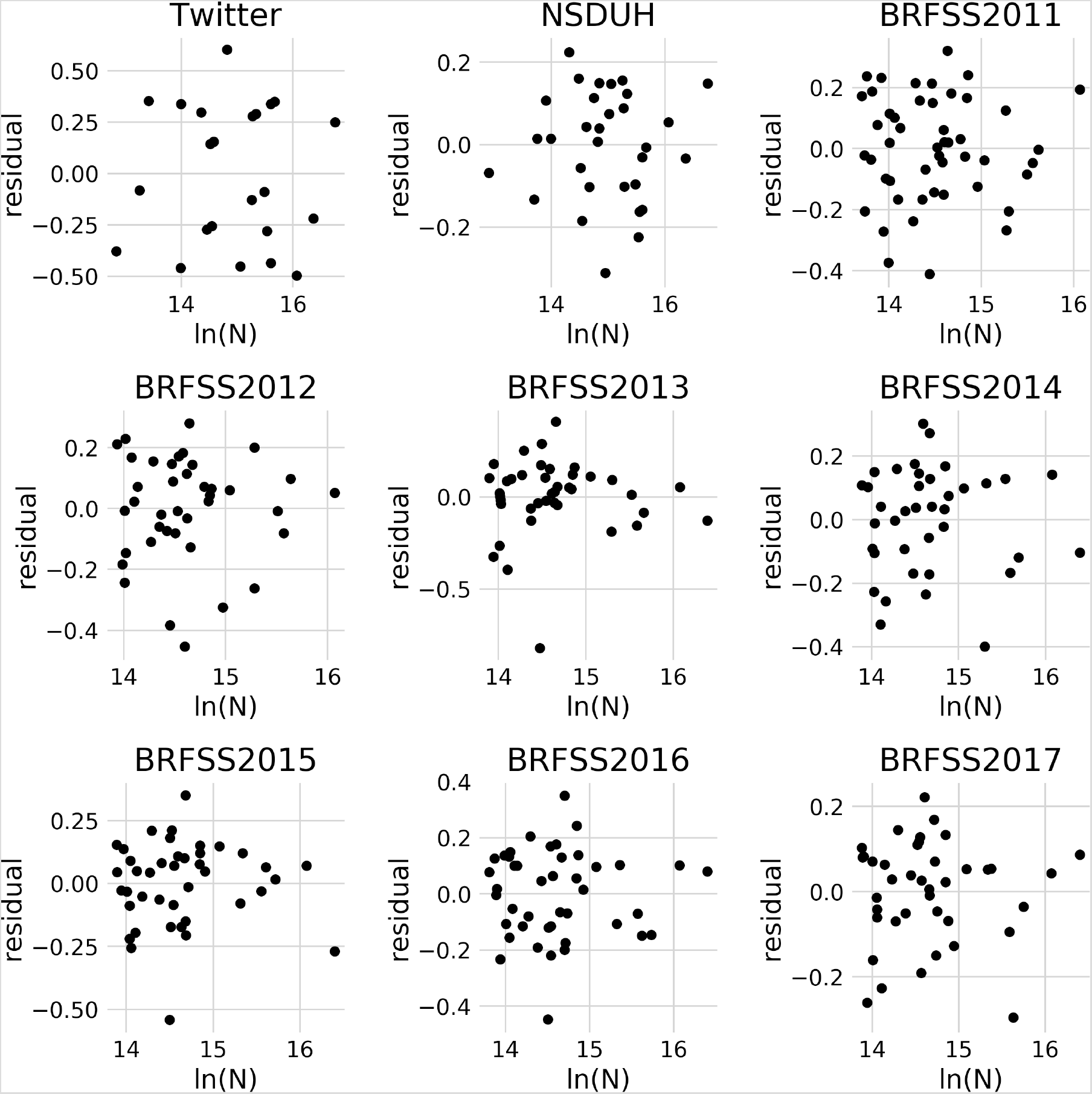
Residuals from OLS models are not correlated with city size. In all datasets, residuals are not correlated with city size (Spearman-r minimum p-value = 0.44). Thus no corrections to estimates of *β* are required.

## Supplementary Tables

**Supplementary Table 1:** MSAs included in the analysis in the main text (Figure 2). Included MSAs are marked with an X.

| MSA | Twitter | BFRSS | NSDUH |
| --- | --- | --- | --- |
| Albuquerque, NM | - | - | X |
| Atlanta-Sandy Springs-Alpharetta, GA | X | X | X |
| Augusta-Richmond County, GA-SC | X | - | - |
| Austin-Round Rock-Georgetown, TX | - | X | - |
| Baltimore-Columbia-Towson, MD | X | X | X |
| Birmingham-Hoover, AL | - | X | - |
| Boston-Cambridge-Newton, MA-NH | X | - | X |
| Buffalo-Cheektowaga, NY | - | X | - |
| Charlotte-Concord-Gastonia, NC-SC | - | X | - |
| Chicago-Naperville-Elgin, IL-IN-WI | X | X | X |
| Cincinnati, OH-KY-IN | X | X | - |
| Cleveland-Elyria, OH | X | X | X |
| Columbus, OH | - | X | - |
| Dallas-Fort Worth-Arlington, TX | X | - | X |
| Denver-Aurora-Lakewood, CO | - | X | X |
| Detroit-Warren-Dearborn, MI | X | - | X |
| Grand Rapids-Kentwood, MI | - | X | - |
| Hartford-East Hartford-Middletown, CT | - | X | - |
| Houston-The Woodlands-Sugar Land, TX | X | X | X |
| Indianapolis-Carmel-Anderson, IN | X | X | - |
| Jacksonville, FL | - | X | - |
| Kansas City, MO-KS | X | X | X |
| Las Vegas-Henderson-Paradise, NV | - | X | X |
| Los Angeles-Long Beach-Anaheim, CA | X | X | X |
| Louisville/Jefferson County, KY-IN | - | X | - |
| Manchester-Nashua, NH | - | - | X |
| Memphis, TN-MS-AR | - | X | - |
| Miami-Fort Lauderdale-Pompano Beach, FL | X | X | X |
| Milwaukee-Waukesha, WI | - | X | - |
| Minneapolis-St. Paul-Bloomington, MN-WI | - | X | X |
| Montgomery, AL | X | - | - |
| Nashville-Davidson–Murfreesboro–Franklin, TN | - | X | X |
| New Orleans-Metairie, LA | X | X | X |
| New York-Newark-Jersey City, NY-NJ-PA | X | - | X |
| Oklahoma City, OK | - | X | - |
| Orlando-Kissimmee-Sanford, FL | X | X | - |
| Philadelphia-Camden-Wilmington, PA-NJ-DE-MD | X | - | X |
| Phoenix-Mesa-Chandler, AZ | - | X | X |
| Pittsburgh, PA | - | X | X |
| Portland-Vancouver-Hillsboro, OR-WA | - | X | X |
| Poughkeepsie-Newburgh-Middletown, NY | X | - | - |
| Providence-Warwick, RI-MA | - | X | - |
| Raleigh-Cary, NC | - | X | - |
| Richmond, VA | X | X | - |
| Riverside-San Bernardino-Ontario, CA | X | X | - |
| Rochester, NY | - | X | - |
| Sacramento-Roseville-Folsom, CA | - | X | - |
| Salt Lake City, UT | - | X | X |
| San Antonio-New Braunfels, TX | - | X | - |
| San Diego-Chula Vista-Carlsbad, CA | - | X | X |
| San Francisco-Oakland-Berkeley, CA | - | X | X |
| San Jose-Sunnyvale-Santa Clara, CA | - | X | - |
| Seattle-Tacoma-Bellevue, WA | X | - | X |
| St. Louis, MO-IL | - | X | X |
| Tampa-St. Petersburg-Clearwater, FL | - | X | X |
| Tulsa, OK | - | - | X |
| Virginia Beach-Norfolk-Newport News, VA-NC | X | X | - |
| Washington-Arlington-Alexandria, DC-VA-MD-WV | - | - | X |

**Supplementary Table 2:** Estimates of the scaling exponent made with BRFSS data from smaller cities that were below the estimated change point for each year.

| Dataset | $\beta$ | 95% CI | $R^2$ | n |
| --- | --- | --- | --- | --- |
| BRFSS2011 | 1.000 | [0.960, 1.039] | 0.952 | 128 |
| BRFSS2012 | 1.001 | [0.961, 1.040] | 0.954 | 122 |
| BRFSS2013 | 1.020 | [0.969, 1.070] | 0.953 | 81 |
| BRFSS2014 | 1.034 | [0.991, 1.077] | 0.969 | 74 |
| BRFSS2015 | 1.044 | [0.996, 1.093] | 0.966 | 67 |
| BRFSS2016 | 0.967 | [0.906, 1.028] | 0.931 | 76 |
| BRFSS2017 | 1.010 | [0.962, 1.058] | 0.959 | 76 |

**Supplementary Table 3:**
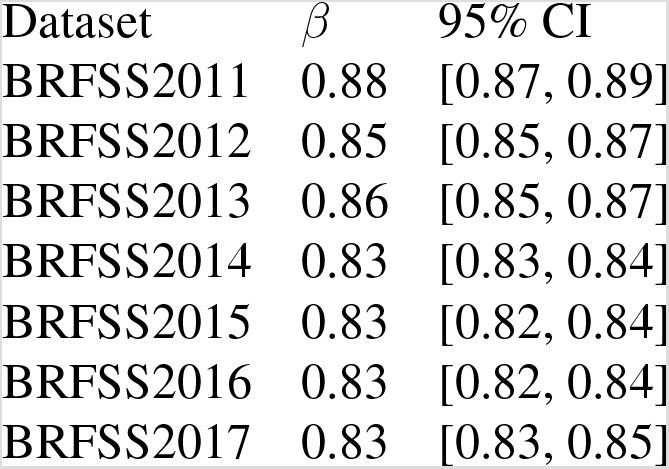
Robustness of scaling exponent estimates made with BRFSS data to variation in the city size below which data was excluded.

**Supplementary Table 4:** Scaling exponent estimates for all BFRSS data. No cities below the change point are excluded.

| Dataset | $\beta$ | 95% CI | $R^2$ | n |
| --- | --- | --- | --- | --- |
| BRFSS2011 | 0.966 | [0.942, 0.991] | 0.974 | 172 |
| BRFSS2012 | 0.956 | [0.931, 0.982] | 0.972 | 161 |
| BRFSS2013 | 0.951 | [0.920, 0.982] | 0.968 | 122 |
| BRFSS2014 | 0.959 | [0.932, 0.987] | 0.978 | 111 |
| BRFSS2015 | 0.961 | [0.932, 0.990] | 0.976 | 109 |
| BRFSS2016 | 0.941 | [0.910, 0.972] | 0.968 | 119 |
| BRFSS2017 | 0.965 | [0.939, 0.991] | 0.980 | 116 |

**Supplementary Table 5:** Robustness of scaling exponent estimates to variation in the minimum number of tweets required for inclusion in the Twitter analyses.

| Minimum Tweets | $\beta$ | 95% CI | # MSAs |
| --- | --- | --- | --- |
| 82 | 0.85 | [0.75, 0.96] | 31 |
| 83 | 0.85 | [0.75, 0.95] | 29 |
| 84 | 0.86 | [0.75, 0.96] | 29 |
| 85 | 0.87 | [0.75, 0.98] | 28 |
| 86 | 0.86 | [0.75, 0.98] | 28 |
| 87 | 0.83 | [0.69, 0.97] | 26 |
| 88 | 0.83 | [0.68, 0.98] | 25 |
| 89 | 0.80 | [0.65, 0.95] | 25 |
| 90 | 0.79 | [0.63, 0.94] | 24 |
| 91 | 0.80 | [0.65, 0.96] | 24 |
| 92 | 0.82 | [0.67, 0.97] | 24 |
| 93 | 0.85 | [0.70, 0.99] | 22 |
| 94 | 0.84 | [0.69, 0.98] | 22 |
| 95 | 0.83 | [0.70, 0.95] | 22 |
| 96 | 0.83 | [0.70, 0.97] | 22 |
| 97 | 0.84 | [0.71, 0.98] | 22 |
| 98 | 0.86 | [0.71, 1.00] | 22 |
| 99 | 0.84 | [0.70, 0.98] | 22 |
| 100 | 0.81 | [0.65, 0.97] | 22 |
| 101 | 0.86 | [0.68, 1.04] | 21 |

**Supplementary Table 6:**
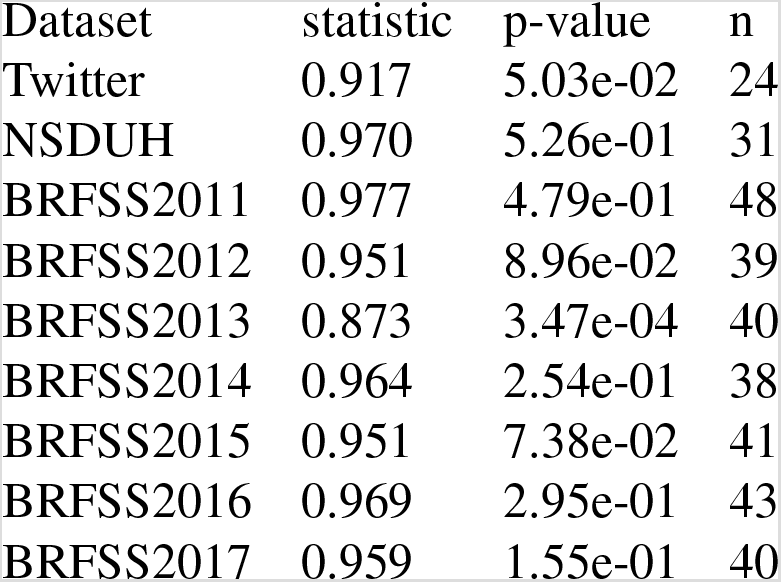
Shapiro-Wilk test of normality on the OLS residuals for each dataset. The residuals from the BRFSS 2013 data fail this normality test due to one outlier city with an negative residual.

| Dataset | statistic | p-value | n |
| --- | --- | --- | --- |
| Twitter | 0.917 | 5.03e-02 | 24 |
| NSDUH | 0.970 | 5.26e-01 | 31 |
| BRFSS2011 | 0.977 | 4.79e-01 | 48 |
| BRFSS2012 | 0.951 | 8.96e-02 | 39 |
| BRFSS2013 | 0.873 | 3.47e-04 | 40 |
| BRFSS2014 | 0.964 | 2.54e-01 | 38 |
| BRFSS2015 | 0.951 | 7.38e-02 | 41 |
| BRFSS2016 | 0.969 | 2.95e-01 | 43 |
| BRFSS2017 | 0.959 | 1.55e-01 | 40 |

## Notes

### Competing Interest Statement

The authors have declared no competing interest.

### Funding Statement

No external funding was recieved

### Author Declarations

Analysis was on publicly available data and as such is covered by the IRB boards of the data publishers

